# MentalQLM: A lightweight large language model for mental healthcare based on instruction tuning and dual LoRA modules

**DOI:** 10.1101/2024.12.29.24319755

**Authors:** Jiayu Shi, Zexiao Wang, Jiandong Zhou, Chengyu Liu, Poly Z.H. Sun, Erying Zhao, Lei Lu

## Abstract

Mental disorders pose significant challenges to healthcare systems and have profound social implications. The rapid development of large language model (LLM) presents new opportunities for improving mental healthcare. However, existing approaches primarily rely on instruction tuning and few-shot in-context learning with massive datasets and large-scale backbone models, leading to significant computational costs. To address these limitations, we propose MentalQLM, a novel lightweight LLM that leverages a dual Low-Rank Adaptation (LoRA) strategy for parameter-efficient fine-tuning. The development of our proposed MentalQLM includes two key stages. Firstly, we perform dataset pruning based on perplexity and diversity analysis to reduce computational load. The first LoRA module is applied during instruction tuning to adapt the base LLM for mental health classification. Secondly, we introduce a dense layer augmented with a second LoRA module, fine-tuned specifically to boost performance on complex multi-class classification problems. Experimental results demonstrate that our proposed MentalQLM, with only 0.5 billion parameters, achieves an average weighted F1-score of 0.778 on mental disorder diagnosis across five benchmark datasets. It outperforms the state-of-the-art instruction-tuned MentaLLaMA-Chat-13B model by 3.2%, and the few-shot tuned GPT-4 model by 17.7%. This promising performance, combined with its significantly lower resource requirements, positions our developed MentalQLM as a cost-effective and efficient solution for real-world mental healthcare applications, especially in computationally constrained environments. Code is available at https://github.com/tortorish/MentalQLM.

## I. Introduction

Mental disorders, including depression, anxiety, and stress, affect one in every eight people globally, posing significant challenges to healthcare systems and carrying substantial social consequences [1], [2]. Text-based dialogue conversations are frequently utilized in mental health interventions [3]. These systems facilitate the application of data-driven computational approaches, which enhance the efficiency of mental healthcare by analyzing various types of textual data, such as clinical notes, patient self-reports, and therapy transcripts. Machine learning techniques play a pivotal role in this context, which demonstrated promising performance in processing and analyzing healthcare data to detect abnormal patterns, sentiments, and early warnings [4], [5]. These capabilities enable precise diagnosis and the development of personalized treatments to improve mental health outcomes.

Traditional machine learning models, such as support vector machine, random forest, and AdaBoost [6], [7], have been used to diagnose different types of mental health conditions. However, these approaches often struggle to handle the complexity and nuanced nature of textual data, as they rely heavily on extensive feature engineering [8]. The advent of deep neural networks has enabled more efficient processing of noisy healthcare data, resulting in more accurate diagnostic outcomes. These models include fully connected neural networks, convolutional neural networks (CNN), and recurrent neural networks (RNN) [9], [10]. Nevertheless, deep learning methods typically require large amounts of labeled data, which presents a significant barrier to their widespread adoption in clinical settings.

Over the past decade, natural language processing (NLP) and computational social science techniques have been increasingly used to detect mental health issues through textual data [11]. These approaches leverage large-scale textual data to identify linguistic patterns, sentiment shifts, and behavioral anomalies associated with mental health conditions [12]. By employing advanced NLP techniques, researchers can analyze unstructured text efficiently, enabling scalable mental health monitoring and personalized care interventions. Pre-trained language models, such as BERT [13], have demonstrated strong performance in stress detection and suicide risk assessment [14]. Furthermore, domain-specific adaptations, such as MentaLLaMA [15] and MentalRoBERTa [16], have further improved accuracy and effectiveness in mental healthcare. Driven by the availability of large-scale datasets and increased computational resources, pre-trained language models, particularly LLM such as ChatGPT [17], LLaMA [18], and Qwen [19], have significantly advanced in both scale and capability. These models have demonstrated promising potential in mental health applications. For instance, ChatGPT has achieved performance comparable to mental health professionals in assessing conditions such as depression and schizophrenia [20], [21]. However, these studies also emphasize the need for continuous refinement to mitigate inherent biases in LLM and improve their clinical reliability. These insights highlight the importance of effective fine-tuning and tailored prompt engineering techniques to enhance the precision and applicability of LLM in mental healthcare.

Instruction fine-tuning and few-shot in-context learning are two effective approaches for adapting LLM to domain-specific tasks [22]. Instruction fine-tuning involves training LLM on curated instruction–output pairs tailored to a target domain [23]. This approach has been proven particularly effective in mental health applications. For example, the Mental-Alpaca and Mental-FLAN-T5 are full-parameter fine-tuned models on mental health classification datasets, and they outperformed GPT-3.5 by a considerable margin and achieved comparable performance to task-specific supervised learning models [24]. Instruction tuning can also be realized using parameter-efficient tuning methods, such as low-rank adaptation (LoRA), which preserve the foundational language capabilities of LLM and increase performance on downstream mental health tasks [25]. In contrast, few-shot prompting enables LLM to perform tasks using a small number of examples within the input prompt [26]. Previous study leveraged in-context learning to adapt LLM for detecting depression in textual data, enabling the model to infer task requirements and generalize effectively to new data [27]. Few-shot prompting is especially useful for scenarios where annotated data is scarce, allowing the model to be efficiently adapted to downstream tasks.

It is noted that most instruction-tuned LLM are typically developed based on backbone models with a large number of parameters [24], which require substantial computational resources for training. While few-shot prompting often leads to long input sequences that significantly increase GPU memory usage and processing time. Moreover, the efficacy of this approach is highly dependent on model size, as it requires models with a substantial scale to achieve satisfactory performance, making this approach less feasible for lightweight applications. Despite the growing interest in adapting LLM to mental healthcare, few studies have focused on developing lightweight and deployable models in this domain. For instance, the smallest model designed for mental health applications, mhGPT, still has 1.98 billion parameters [25], which poses challenges for efficient deployment. Therefore, it is critical to investigate how factors such as model size and quantization techniques affect the performance and usability of LLM in resource-constrained environments.

The development of lightweight LLM, or on-device LLM, is crucial for improving the accessibility and efficiency of mental healthcare, particularly in resource-constrained settings. For instance, low-income developing countries (LIDCs) face a growing mental health burden but have limited computational resources and infrastructure [28]. These regions urgently need resource-efficient solutions capable of delivering effective mental health support. Deploying lightweight LLM on edge devices such as mobile phones enables delivering personalized, ubiquitous psychological support, offering scalable and cost-effective solutions for mental healthcare in real-world settings [29]. Additionally, such models are valuable in time-sensitive scenarios, supporting early diagnosis and intervention by detecting subtle linguistic cues associated with mental disorders [30].

Furthermore, as illustrated in Figure 1, a collaborative framework can be established to leverage the strengths of both lightweight edge LLM and large-scale cloud LLM [31]. Edge LLM, deployed on personal devices, can provide timely mental health suggestions and record therapeutic dialogues. These interactions can be stored as electronic health records (EHRs) and sent to cloud-based LLM for more comprehensive analysis. Meanwhile, advanced techniques such as knowledge distillation can be applied to transfer insights from the cloud models back to the edge models, enhancing their performance while preserving computational efficiency [32]. By enabling lightweight LLM to approach the capabilities of their larger counterparts, this collaborative approach can help close gaps in mental health service delivery, offering real-time, personalized support and promoting the widespread adoption of AI-driven mental healthcare solutions. However, a major limitation of lightweight LLM remains their reduced parameter scale, which often results in diminished performance in care delivery tasks [33]. Therefore, there is a pressing need to design lightweight LLM that can operate efficiently on resource-constrained devices while still delivering accurate, timely, and clinically useful diagnostic support.

**Fig. 1:**
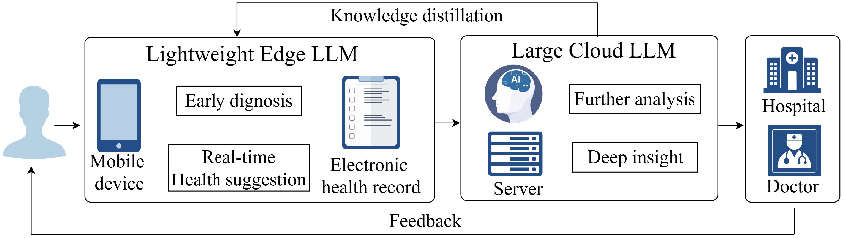
Collaboration between edge LLM and cloud LLM.

In this paper, we propose a novel framework for developing lightweight LLM tailored for mental healthcare applications. The model, named **Mental**-**Q**wen-**L**ightweight language **M**odel (MentalQLM), is built on the Qwen1.5-0.5B-Chat backbone [19]. To enhance the model’s performance on mental health tasks, we introduce a new dual LoRA strategy, which optimizes the lightweight LLM for improving efficiency and task-specific adaptability. The first LoRA module focuses on instruction tuning, enabling the model to adapt effectively to downstream mental health tasks by leveraging task-specific instruction-output pairs. The second LoRA module is designed to enhance the model’s multi-class classification capabilities, making it suitable for handling complex mental health tasks. This dual strategy combines the benefits of instruction tuning and traditional supervised learning, offering a comprehensive approach to model optimization. In addition, knowledge from the large LLM is transferred to MentalQLM through instruction tuning, enabling the lightweight model to achieve performance comparable to its large-scale counterparts.

To the best of our knowledge, this is the first study that comprehensively explores the instruction-following capabilities of models with varying scales and quantization methods in the context of mental health applications. By addressing existing limitations, this work aims to advance the development of lightweight LLM, allowing for more accessible and efficient mental healthcare delivery. The main contributions of this paper are summarized as follows.

- A novel lightweight large language model MentalQLM is proposed for the diagnosis and reasoning of mental health conditions.
- A new dual LoRA strategy is developed to optimize the lightweight MentalQLM for improving the efficiency and task-specific mental health applications.
- Extensive experiments are conducted on six benchmark mental health datasets and multilingual evaluation to validate the effectiveness of the proposed MentalQLM.
- Compared to state-of-the-art LLM (e.g., GPT-4 and MentaLLaMA), the MentalQLM has only 0.5 billion parameters yet achieves superior or comparable performance in mental disorder diagnosis and reasoning tasks.

The remainder of this paper is organized as follows. Section II presents the development of our proposed MentalQLM, and Section III shows experiments and results of using MentalQLM for mental disorder diagnosis and reasoning. Section IV discusses the strengths and limitations of this study. Finally, conclusions are drawn in Section V.

## II. Methodology

This study proposes MentalQLM, a novel lightweight LLM designed for mental healthcare applications. The development process includes two key stages: a parameter-efficient instruction tuning stage and a multi-class classification fine-tuning stage. Figure 2 illustrates an overview of the methodology for developing the lightweight LLM using dual LoRA modules. In the first stage, instruction-output pairs are selected based on quality and diversity assessments. These pairs are categorized into binary classification datasets and multi-class classification datasets. The first LoRA module is then instruction-tuned using these datasets, optimized with the standard language modeling loss function. In the second stage, the multi-class fine-tuning process further refines the model. The prompts used in the instruction-tuning phase are removed, as they are primarily beneficial for training the initial LLM. Retaining prompts in the fine-tuning stage would also increase computational overhead unnecessarily. The second LoRA module is fine-tuned using the categorical cross-entropy loss function, enhancing the performance of MentalQLM on complex multiclass classification tasks. Detailed procedures of developing the MentalQLM are presented in Figure 3 and the following sections.

**Fig. 2:**
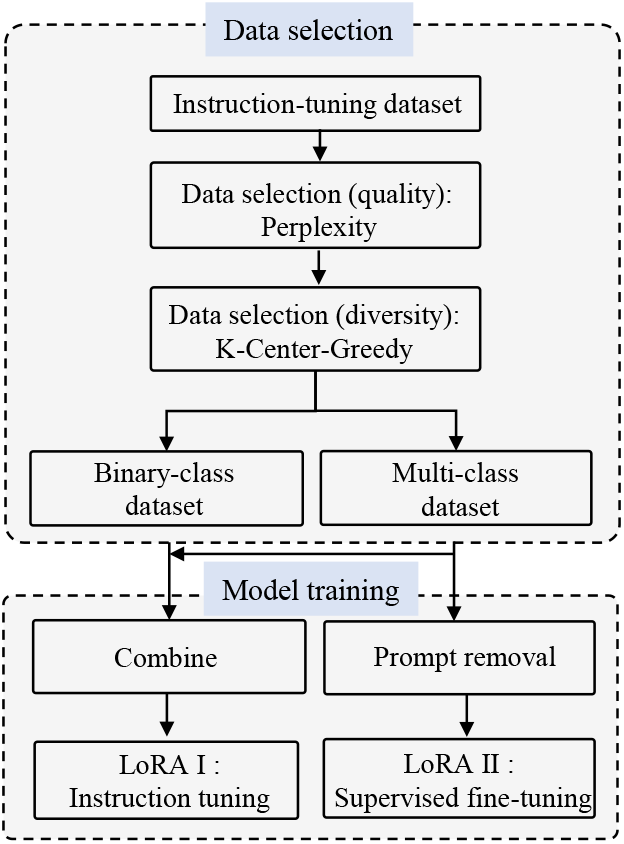
Flowchart of our proposed fine-tuning methodology.

**Fig. 3:**
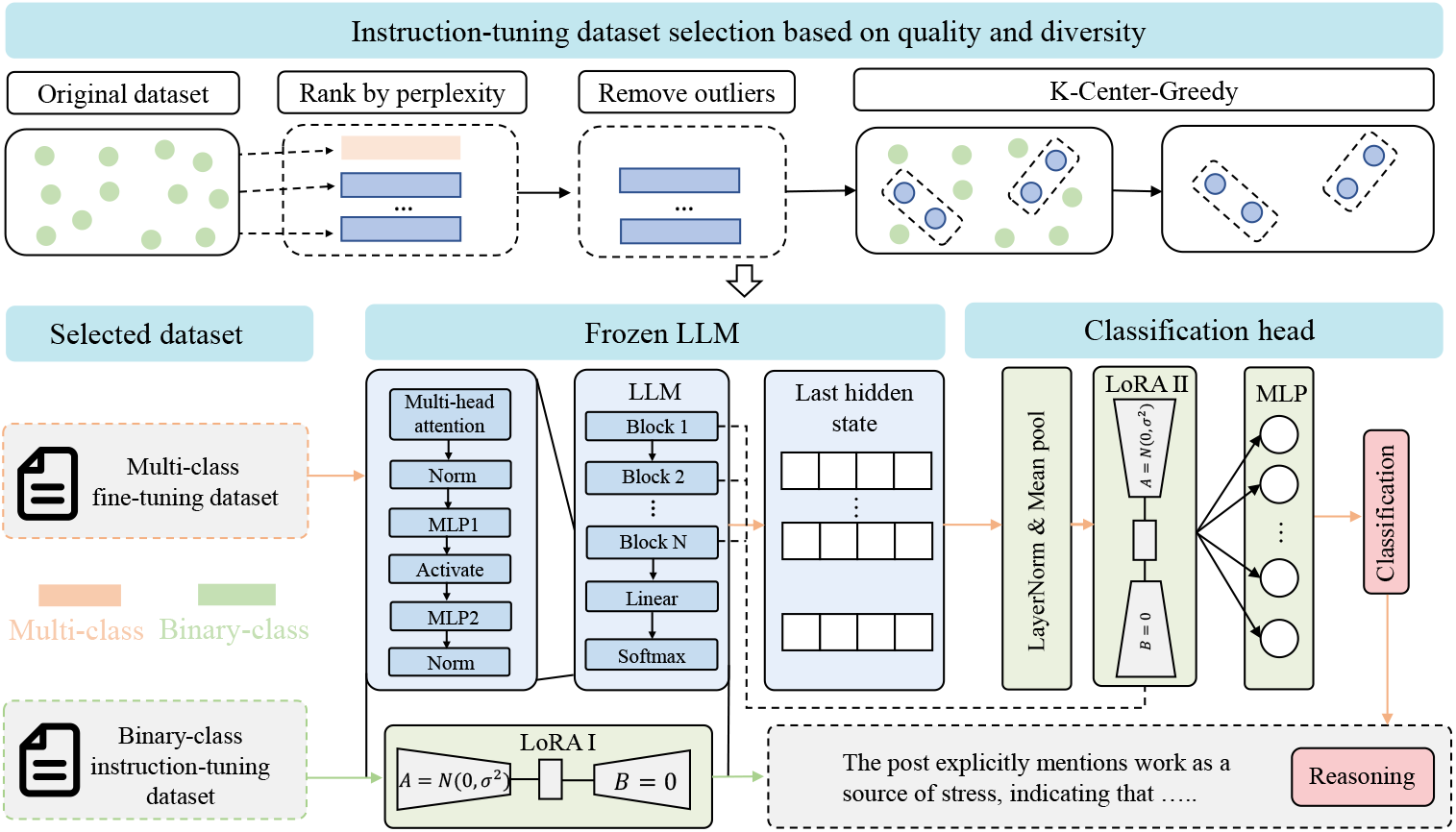
Detailed pipeline of developing our proposed MentalQLM for mental health diagnosis and reasoning. (1) Data selection: Instruction-output pairs are selected based on quality and diversity assessments. (2) Frozen LLM and training: the first LoRA module is instruction-tuned using the binary class instruction-tuning dataset. (3) Classification: an additional classification head is added, and the second LoRA module is used for fine-tuning with the multi-class classification dataset.

## A. Data Pruning by Perplexity and Diversity Analysis

Previous research indicated that a small amount of high-quality data during instruction tuning can achieve performance comparable to models trained on much larger datasets for downstream tasks [34]. Building on this insight, we propose a data selection method that prioritizes quality and diversity. This approach ensures that the selected instruction-output pairs effectively represent the target domain while significantly reducing computational demands without compromising accuracy [35].

As illustrated in the upper part of Figure 3, data points in the original dataset are ranked by perplexity, where the perplexity is computed based on the untuned backbone model for each instruction-output pair. Outliers with high perplexity are eliminated to ensure quality of the dataset. Only data points within the perplexity range [*Q*_1_ −1.5· *IQR, Q*_3_ + 1.5 ·*IQR*] are kept, which can be formalized as

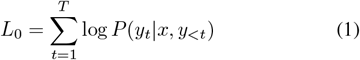

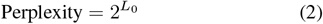

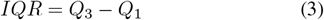

where *x* denotes the instruction, *y* denotes the output sentence, *P* denotes the probability value for generating token *y*_*t*_, given the instuction *x* and previous tokens *y*_*<t*_, *Q*_1_ and *Q*_3_ are the first quartile and third quartile of all perplexity values respectively, and *IQR* denotes the interquartile range.

Next, the K-Center-Greedy approach is utilized to select a diverse subset of data for instruction fine-tuning. This algorithm ensures comprehensive coverage across the feature space while minimizing redundancy and maximizing information density. As outlined in Algorithm 1, the process begins by determining the value of *K*, where *K* = *p* ·|*d*|, *p* denotes the proportion with a default value of 0.25, and *d* denotes the number of samples in each dataset. An initial center *e* is randomly selected from the dataset. Then, each new center is chosen to maximize the minimum distance *d*(*e, c*) from any existing center, ensuring diverse representation throughout the dataset. This iterative process continues until *K* centers are established, effectively achieving a balance between coverage and compactness.

### Algorithm 1

*K*-Center-Greedy Algorithm.

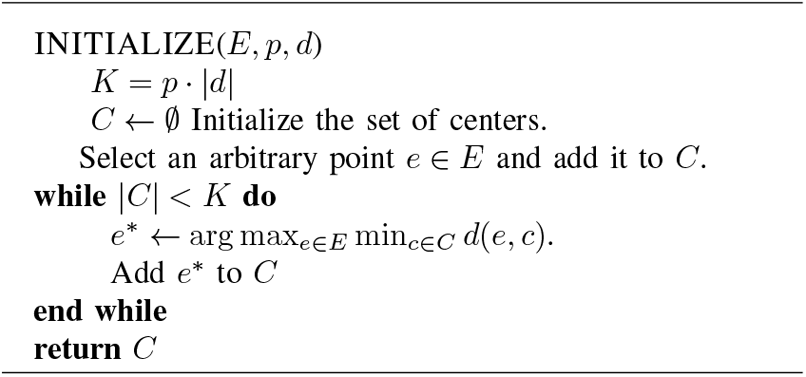

### B. Parameter Efficient Instruction Tuning

We adopted a parameter-efficient fine-tuning (PEFT) approach to implement the instruction tuning of LLM, where only a small subset of parameters is updated. This approach significantly reduces computational costs, preserves the foundational capabilities of the LLM, and prevents catastrophic forgetting of previously acquired knowledge. Instruction tuning ensures that the model adapts effectively to new tasks while maintaining its original knowledge base. Specifically, MentalQLM is instruction-tuned using the LoRA method [36], which updates only a low-rank approximation of the weight matrices. This method can be formalized as follows [36],

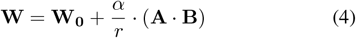

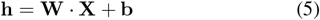

where **W**_**0**_ ∈ ℝ^*m×n*^ denotes the original matrix, **A** ∈ ℝ^*m×r*^ and **B** ∈ ℝ^*r×n*^ denotes two smaller matrices, *r* ≪ min(*m, n*), *α* is a scaling coefficient that adjusts the contribution of the low-rank matrices **A** and **B** to the desired weights **W, h** denotes the output hidden state, and **b** is the biased term.

During the instruction-tuning stage, the LoRA method is applied to all linear layers to optimize performance across a wide range of tasks. By leveraging the low-rank approximation, this approach effectively reduces computational costs while maintaining the flexibility and adaptability of the model. The training objective is to minimize the negative log-likelihood, which measures the discrepancy between the predicted token probabilities and the actual ground-truth tokens. For an instruction-output pair (*x, y*), the loss *L*_1_ is defined as

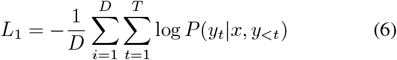

where *D* denotes the collection of instruction-output pairs, the sequence *y* is made up of *T* tokens, *P* (*y*_*t*_|*x, y*_*<t*_) is the probability of generating token *y*_*t*_ given the instruction *x* and previous tokens *y*_*<t*_. Note that only loss of the output sentence is computed in the instruction-tuning stage.

### C. Multi-class Classification Fine-tuning

To enhance the ability of MentalQLM on complex multi-class classification tasks, we proposed a fine-tuning method. Compared with traditional text-embedding models such as BERT [13], and RoBERTa [37], it is difficult to fine-tune LLM on downstream classification tasks. In addition, the model is prone to overfitting on the training set due to the adjustment of a large number of parameters during fine-tuning, which also results in significant computational costs.

To address this issue, we introduce a second LoRA module, which is appended exclusively to the queries and values of the multi-head attention mechanism, as indicated in Eq. (7). This design ensures that the second LoRA module (LoRA II) is much smaller than the first LoRA module (LoRA I) to reduce the parameter count.

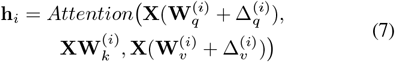

where **X** denotes the input vector,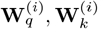 and 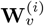 denote the query, key, and value matrix of the *i*th transformer decoder block, *Attention*(·) is the operator of multi-head attention, 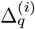, and 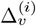 denotes the LoRA matrices of 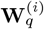 and 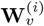.

Next, the whole dataset is fed into LLM to get sentence embedding, and all parameters in LLM are frozen as illustrated in Figure 3. It is noted that pretrained language models, such as BERT [13], utilize a [*CLS*] token as a compressed sentence representation for classification tasks. However, self-regressive unidirectional models, such as Qwen, do not include this token [19]. Therefore, we use the final hidden states from the last decoder block as the sentence representation, and mean pooling is applied across the sequence length dimension. Suppose an input sentence **S** is tokenized into (**w**_**1**_, **w**_**2**_, …, **w**_**n**_), each token **w**_**i**_ corresponds to an embedding vector **e**_**i**_ ∈ *R*^*d*^, *n* is length of the sentence, and *d* denotes length of the embedding. Then, a one-dimensional vector **h** ∈ *R*^*d*^ can be obtained through mean pooling. To enhance computational efficiency and convergence speed, layer norm is utilized to process the sentence embeddings. Finally, a dense layer is appended to the model to regress the dimension to the number of classes. We use the categorical cross-entropy loss in the fine-tuning stage, and it can be formulated as:

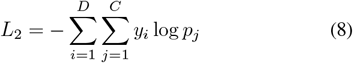

where *C* is the number of classes, *y*_*i*_ is 1 if the sample belongs to class *i*, and *p*_*j*_ is the predicted probability of class *j*, and *D* denotes the collection of instruction-output pair. This loss function encourages the model to assign high probabilities to the correct class labels while minimizing those for incorrect categories. During the multi-class fine-tuning stage, parameters in the LoRA matrices 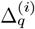 and 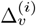 are updated with parameters in the dense layer using the categorical cross-entropy loss. This approach ensures effective adaptation to complex classification tasks while maintaining parameter efficiency.

Based on the classification results, the reasoning module of MentalQLM generates an explanation for the predicted label. This approach combines the strengths of supervised learning for mental disorder classification with the powerful text generation capabilities of LLM, ensuring both precise classification and detailed reasoning. To summarize, for binary classification tasks, MentalQLM simultaneously generates classification labels and reasoning sentences; While for multi-class classification tasks, MentalQLM first predicts the classification label using the classification head, then generates a reasoning sentence based on the prediction, providing explanations tailored to the specific class.

## III. Experiments AND Results

### A. Datasets and Experiment Settings

We benchmark our proposed model on the Interpretable Mental Health Instruction (IMHI) dataset in English [15], which includes 105K data samples supporting multi-task and multi-source LLM instruction tuning and evaluation. In particular, five mental health IMHI sub-datasets are used to validate our developed MentalQLM, including the DR, Dreaddit, Irf, MultiWD, and SAD datasets [15]. All these data are converted to question-answer pairs to meet the requirement of instruction tuning, and they are categorized as binary-class classification datasets and multi-class classification datasets.

In addition to the above monolingual datasets, we evaluate the MentalQLM on multilingual datasets, following prior research by using the DEP-SEVERITY dataset in English along with its translations in six other languages [38], [39]. The dataset comprises Reddit posts and is constructed based on clinical standards for diagnosing depression. DEP-SEVERITY is an imbalanced, multi-class classification dataset designed to assess the severity of depression in users, with categories ranging from minimal, mild, and moderate to severe levels.

Detailed description of these datasets and the selection of model hyperparameters can be found in the Supplementary Material (Appendix I).

### B. Results of Classification Performance

#### 1) Monolingual Evaluation

Experiments are carried out to compare the performance of the state-of-the-art (SOTA) models in terms of classification ability. We benchmarked classification performance using both in-context learning models and instruction-tuned models.

- In-context learning models: They include zero-shot and few-shot models, consisting of GPT-3.5 and GPT-4. Instructions are combined with format requirements to make the LLM output the classification label. To fully utilize the strong learning ability of large-scale LLM, few-shot examples are given for demonstration.
- Instruction-tuning models: The instruction-tuning models consist of MentaLLaMA-7B, MentaLLaMA-7B-Chat and MentaLLaMA-13B-Chat [15], which are instruction-tuned on the LLaMA backbone. Instruction fine-tuning can precisely calibrate models with domain knowledge, improving the classification ability and reasoning ability of LLM on downstream tasks.

As shown in Table I, the performance of our proposed MentalQLM is benchmarked against various LLM across five datasets. Despite its reduced model size, MentalQLM achieves a higher average classification accuracy compared to its larger counterparts. Notably, it outperforms MentaLLaMA-13B-Chat on the Dreaddit and MultiWD datasets by 9.32% and 3.69%, respectively. In contrast, in-context learning models such as GPT-3.5 and GPT-4 exhibit strong zero-shot performance on simpler tasks, such as stress and depression detection on the DR dataset. However, their performance drops significantly on challenging tasks, such as those in the Irf and MultiWD datasets. This highlights the importance of instruction-tuning for specific downstream tasks.

**TABLE I:**
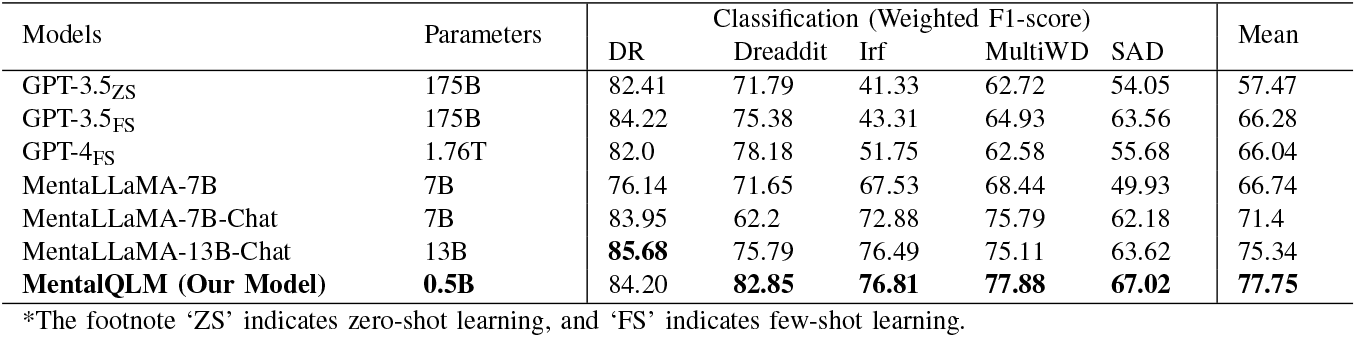
Monolingual classification performance of MentalQLM and comparison with SOTA LLM.

| Models | Parameters | Classification (Weighted F1-score) |  |  |  |  | Mean |
| --- | --- | --- | --- | --- | --- | --- | --- |
|  |  | DR | Dreaddit | Irf | MultiWD | SAD |  |
| GPT-3.5<sub>ZS</sub> | 175B | 82.41 | 71.79 | 41.33 | 62.72 | 54.05 | 57.47 |
| GPT-3.5<sub>FS</sub> | 175B | 84.22 | 75.38 | 43.31 | 64.93 | 63.56 | 66.28 |
| GPT-4<sub>FS</sub> | 1.76T | 82.0 | 78.18 | 51.75 | 62.58 | 55.68 | 66.04 |
| MentaLLaMA-7B | 7B | 76.14 | 71.65 | 67.53 | 68.44 | 49.93 | 66.74 |
| MentaLLaMA-7B-Chat | 7B | 83.95 | 62.2 | 72.88 | 75.79 | 62.18 | 71.4 |
| MentaLLaMA-13B-Chat | 13B | **85.68** | 75.79 | 76.49 | 75.11 | 63.62 | 75.34 |
| **MentalQLM (Our Model)** | **0.5B** | 84.20 | **82.85** | **76.81** | **77.88** | **67.02** | **77.75** |

\*The footnote ‘ZS’ indicates zero-shot learning, and ‘FS’ indicates few-shot learning.

For the multi-class SAD dataset, we use random up-sampling techniques to address the class imbalance issue [40]. Specifically, this process randomly duplicates samples from minority classes to balance the class distribution before training. It involves supplementing the minority classes with additional, replicated instances to match the number of samples in the majority class. It can be seen from Table I that both the in-context learning GPT models and instruction-tuned MentaLLaMA models have reduced classification performance on the multi-class SAD dataset. In contrast, our proposed MentalQLM achieves better classification performance by using task-specific fine-tuning, which incorporates an additional classification head consisting of the second LoRA module and a dense layer. During the fine-tuning stage, parameters of the base LLM are frozen to preserve its strong foundational language capabilities. Only parameters of the classification head are updated during backpropagation, making the process efficient and focused. The results in Table I show that this fine-tuning approach significantly improves classification accuracy on the SAD dataset, enabling more precise diagnosis of multi-class mental health disorders.

We also compute the p-value between the diagnostic results from MentaLLaMA-7B-Chat and MentalQLM using the bootstrap approach [41], [42]. We performed 1,000 resamplings on each of these five datasets, and calculated the p-values with 0.56 on DR, 0.84 on Dreaddit, 0.61 on Irf, 0.80 on MultiWD, and 0.52 on SAD. These values indicate that our proposed MentalQLM achieves comparable classification accuracy to the much larger MentaLLaMA-7B-Chat model.

#### 2) Multilingual Evaluation

We additionally evaluate MentalQLM for the classification of mental disorders on the DEP-SEVERITY dataset, and its translations in six languages, including Turkish, French, Portuguese, German, Greek, and Finnish [38], [39]. We note that the DEP-SEVERITY dataset is imbalanced, and we applied the random up-sampling technique to the mild, moderate, and severe classes, equating their sample sizes to that of the minimum class [40]. We benchmarked the multilingual classification performance of our proposed MentalQLM against SOTA models as reported in previous research [38], including the GPT-4o-mini with zero-shot learning, Llama3.1 with zero-shot learning, GPT-3.5-turbo with zero-shot learning, and GPT-3.5 with one-shot learning. To ensure consistency with their evaluation, we reported the macro-averaged precision, recall, and F1 scores for each class and language [38]. The multilingual classification performance of our proposed MentalQLM almost outperforms all these SOTA models, as an example, it can be seen from Table II that our proposed MentalQLM has higher F1-scores than GPT-4o-mini zero-shot learning across all the multilingual datasets. The results in Table II indicate that targeted fine-tuning enhances task-specific performance and stability, therefore substantiating the effectiveness of our MentalQLM in multilingual scenarios.

**TABLE II:** Multilingual classification performance of MentalQLM and comparison study.

| Categories | English |  |  | Turkish |  |  | French |  |  | Portuguese |  |  | German |  |  | Greek |  |  | Finnish |  |  |
| --- | --- | --- | --- | --- | --- | --- | --- | --- | --- | --- | --- | --- | --- | --- | --- | --- | --- | --- | --- | --- | --- |
|  | Pr | Rec | F1 | Pr | Rec | F1 | Pr | Rec | F1 | Pr | Rec | F1 | Pr | Rec | F1 | Pr | Rec | F1 | Pr | Rec | F1 |
| Minimum | 87.1 | 93.4 | 90.1 | 82.4 | 88.8 | 85.5 | 87.7 | 88.0 | 87.9 | 86.2 | 84.6 | 85.4 | 85.9 | 79.9 | 82.8 | 78.7 | 72.6 | 75.5 | 79.5 | 85.3 | 82.3 |
| Mild | 56.5 | 44.8 | 50.0 | 37.5 | 31.0 | 34.0 | 60.0 | 41.4 | 49.0 | 25.0 | 31.0 | 27.7 | 34.6 | 31.0 | 32.7 | 13.2 | 17.2 | 14.9 | 19.1 | 13.8 | 16.0 |
| Moderate | 53.1 | 42.5 | 47.2 | 28.6 | 25.0 | 26.7 | 37.5 | 52.5 | 43.8 | 37.5 | 45.0 | 40.9 | 30.8 | 30.0 | 30.4 | 18.2 | 15.0 | 16.4 | 32.1 | 22.5 | 26.5 |
| Severe | 43.5 | 35.7 | 39.2 | 33.3 | 21.4 | 26.1 | 40.0 | 28.6 | 33.3 | 22.2 | 14.3 | 17.4 | 20.0 | 35.7 | 25.6 | 8.7 | 14.3 | 10.8 | 13.8 | 14.3 | 14.0 |
| MentalQLM | 60.0 | 54.1 | **56.6** | 45.5 | 41.6 | **43.1** | 56.3 | 52.6 | **53.5** | 42.7 | 43.7 | **42.8** | 42.8 | 44.2 | **42.9** | 29.7 | 29.8 | **29.4** | 36.1 | 34.0 | **34.7** |
| GPT-4o-mini [38] | 35.0 | 35.0 | 27.0 | 33.0 | 31.0 | 21.0 | 34.0 | 34.0 | 26.0 | 34.0 | 36.0 | 27.0 | 34.0 | 35.0 | 27.0 | 34.0 | 33.0 | 24.0 | 34.0 | 33.0 | 24.0 |

\*Pr indicates precision, Rec indicates recall.

### C. Performance of Reasoning Ability

We investigated the reasoning performance of our developed MentalQLM across different metrics, including the widely used BLEU-4 score, BERT-score, and BART-score [43]. In addition, recent studies suggested that evaluations conducted by well-trained LLM align closely with human assessments [20], [21]. For example, LLM such as ChatGPT are increasingly recognized for their effectiveness in text evaluation, offering nuanced and reliable judgments [21]. Motivated by findings in these studies, in addition to the BLEU-4 score, BERT-score, and BART-score, we also employ GPT-4o to rate the quality of reasoning sentences [44]. The prompt template used for evaluation is detailed in the Supplementary Material (Appendix IV).

For each of the five datasets, 200 instruction-output pairs were randomly selected for evaluation, resulting in a total of 1,000 pairs. As shown in Figure 4, we compared the evaluation scores of sentences generated by MentalQLM and MentaLLaMA-7B-Chat.

**Fig. 4:**
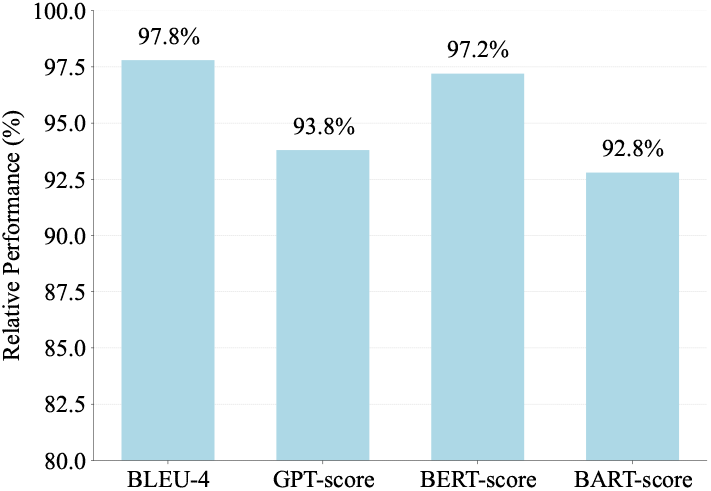
Reasoning performance analysis of MentalQLM.

Using the scores of MentaLLaMA-7B-Chat as the 100% baseline, the results as shown in Figure 4 indicate that our developed MentalQLM, with only 7% of the parameters and 25% of the training data, achieves 95.4% of the inference performance on average across four metrics. Notably, on the semantic similarity-based metric BERT-Score, the model achieves 97.2%, closely approximating the much larger MentaLLaMA-7B-Chat, further validating the high quality of the sentences generated by MentalQLM.

We provide three examples of reasoning sentences generated by MentalQLM and MentaLLaMA-7B-Chat, along with the GPT-4o scores and prompts, in the Supplementary Material (Appendix VII). In particular, the input prompt to GPT-4o evaluates four criteria, i.e., helpfulness, relevance, accuracy, and detail level, with scores ranging from 1 to 10 for each sample [44]. These criteria ensure unbiased and comprehensive feedback.

### D. Impacts of Data Size and Data Quality

As described in Section II, the datasets used for training the MentalQLM model are pruned at two dimensions: data quality and data diversity. At the data quality level, we use perplexity to filter the dataset. Perplexity is a key metric in language modeling that quantifies model’s predictive capability by measuring how well it assigns probabilities to a sequence of text. Lower perplexity indicates greater confidence and better predictive performance, especially on unseen data. At the data diversity level, we apply the K-Center-Greedy algorithm to ensure the maximal coverage of sample space while with the minimal number of representative points. In the following, we perform ablation studies to quantitatively analyze the impact of our developed data trimming techniques.

#### 1) Data Size Analysis

The relationship between dataset size and model performance is analyzed and illustrated in Figure 5. When the dataset size increases from 0% to 3%, there is a rapid rise in both the BLEU-4 score and the weighted F1 score. This indicates that even a small subset of data can significantly enhance the model’s prediction performance. When the dataset size increases from 3% to 25%, the scores continue to improve gradually. The curves of incremental improvement suggest that while additional data further enhances the model performance, the rate of improvement begins to diminish. Beyond the ratio of 25%, the score remains stable, indicating a minimal impact on performance with larger dataset sizes. These results suggest that 25% of the dataset is generally sufficient to achieve near-optimal performance.

**Fig. 5:**
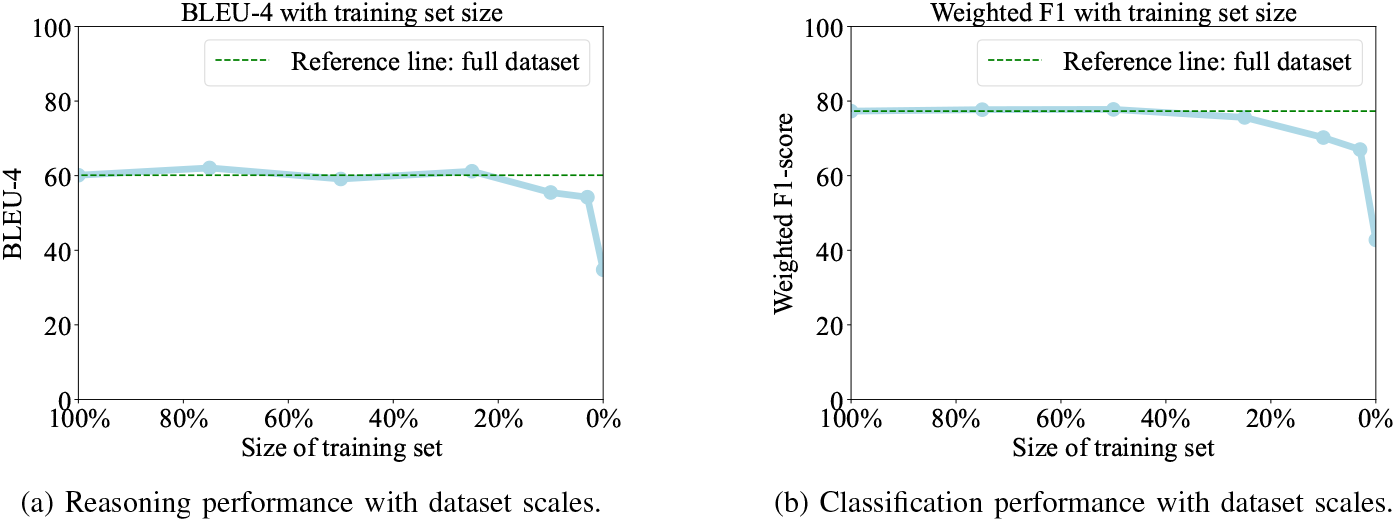
Classification and reasoning performance with different data sizes.

#### 2) Data Quality Analysis

We also investigated the influence of data quality on model performance. Figure 6 (a) illustrates the perplexity distribution of the instruction-output pairs across the five datasets, where outliers with extremely high perplexity are removed to ensure dataset quality. Figure 6 (b) shows results of an ablation study conducted on subsets comprising 3% and 10% of the entire dataset. When only 3% of the data is used, outliers have an adverse impact on model performance. Specifically, if the outliers are included during training, the BLEU-4 score has a significant drop. This finding underscores the critical importance of maintaining high quality data when training resources are constrained or limited samples are available. In contrast, with 10% of the data, the negative effect of outliers is mitigated due to the introduction of additional instruction-output pairs.

**Fig. 6:**
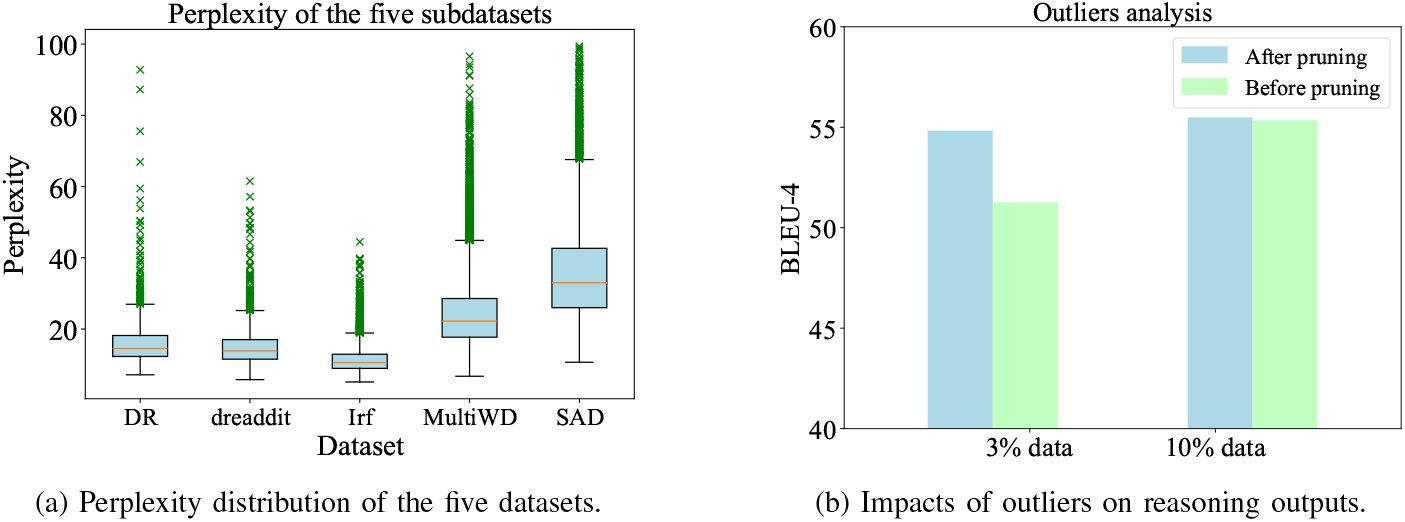
Perplexity distribution and data quality analysis.

In the Supplementary Material (Appendix VI), examples of instruction-output pairs with high perplexity are presented, which are ambiguous for mental disorder. These examples will interfere with the model’s ability to learn the correct reasoning path. These findings highlight that data quality, rather than sheer quantity, is a pivotal factor in ensuring model efficacy, particularly in data-restricted scenarios.

### E. Analysis Abnormal Characters

Hallucination is a common limitation of LLM, where they may generate unsubstantiated or irrelevant content during reasoning [45]. As shown in Figure 7, we identified 259 Chinese characters among the 613,953 words in the reasoning text generated by MentalQLM, accounting for just 0.0004 of the total words. A similar phenomenon is observed in MentaLLaMA-7B-Chat, which contains approximately 0.0005 non-English characters in its reasoning outputs. To further investigate this issue, we compared the generation of abnormal characters across LLM of different scales. As illustrated in Figure 7, replacing the backbone with Qwen1.5-7B-Chat significantly increased the number of Chinese characters. This highlights the advantage of our fine-tuned lightweight MentalQLM in producing more accurate and focused predictions.

**Fig. 7:**
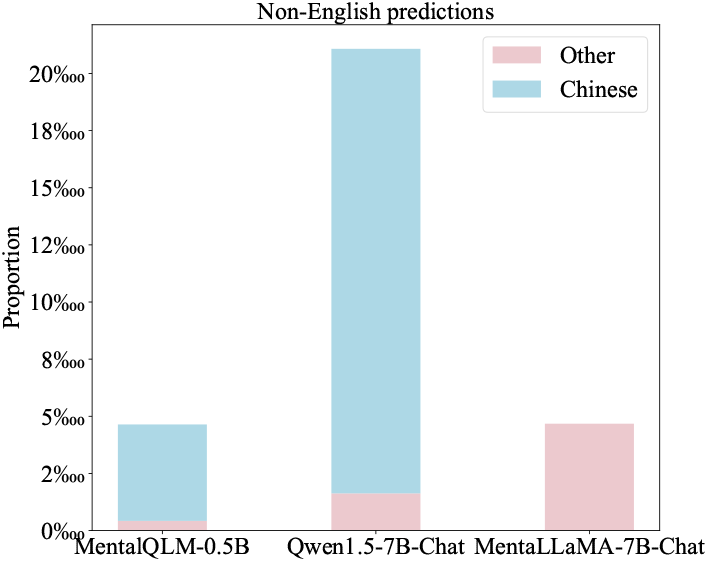
Non-English predictions from different LLM.

The detailed examples of non-English characters in the generated text are presented in the Supplementary Material (Appendix V).

### F. Ability of Instruction Following and Embedding Visualization

The section explores the relationship between the model size and instruction-following ability, and also investigates text embeddings of LLM for various downstream tasks without task-specific pre-training.

#### 1) Instruction-Following Ability of Models with Different Sizes

The performance of models with different parameter scales are presented in Table III. We fine-tuned all these different models using the first LoRA module for comparison. It can be seen from Table III that instruction tuning demonstrates effectiveness for models of various sizes. While large-scale models are generally recognized for their superior capabilities, smaller models can achieve comparable performance when instruction-tuned. As demonstrated in Table III, the classification and reasoning abilities of the lightweight Qwen1.5-0.5B-Chat model are on par with those of larger models. Therefore, we selected Qwen1.5-0.5B-Chat as backbone for designing our lightweight LLM.

**TABLE III:** Instruction-following ability of models with different parameter scales.

| Models | Classification (Weighted F1-score) |  |  |  |  | Reasoning (BLEU-4 score) |  |  |  |  |
| --- | --- | --- | --- | --- | --- | --- | --- | --- | --- | --- |
|  | DR | Dreaddit | Irf | MultiWD | SAD | DR | Dreaddit | Irf | MultiWD | SAD |
| Qwen2-7B-Chat | 77.88 | 81.64 | 75.63 | 76.73 | 64.33 | 62.58 | 60.32 | 61.98 | 65.81 | 65.30 |
| Qwen1.5-7B-Chat | 86.86 | 86.45 | 78.79 | 79.41 | 59.27 | 61.82 | 61.20 | 65.15 | 64.98 | 62.65 |
| Qwen1.5-4B-Chat | 86.17 | 84.06 | 76.01 | 78.82 | 62.57 | 62.05 | 60.48 | 61.41 | 65.35 | 63.95 |
| Qwen1.5-1.8B-Chat | 84.44 | 84.54 | 77.90 | 78.37 | 63.45 | 61.86 | 60.42 | 61.39 | 66.15 | 64.45 |
| Qwen1.5-0.5B-Chat | 82.38 | 80.91 | 76.76 | 78.75 | 58.31 | 60.81 | 58.70 | 61.04 | 64.69 | 63.37 |

The results in Table III also showed that Qwen1.5-7B-Chat consistently outperforms its counterpart, Qwen2-7B-Chat, on most datasets. This suggests that instruction tuning outcomes have a limited correlation with adjacent model versions, highlighting the importance of task-specific evaluation. However, on the multi-class classification dataset SAD, the weighted F1 scores across all models are relatively low, particularly for the 0.5B parameter-scale model. This underscores the need to enhance the classification capabilities of lightweight LLM for complex multi-class classification tasks.

As the models presented in Table III are fine-tuned using only one LoRA module. This setup provides an opportunity to compare the performance of a single LoRA module with our proposed dual LoRA module. Specifically, the Qwen1.5-0.5B-Chat model, fine-tuned with a single LoRA module, achieves a diagnostic accuracy of 58.31 on the multi-class SAD dataset (Table III). In contrast, our proposed MentalQLM, which uses Qwen1.5-0.5B-Chat as the backbone and the dual LoRA strategy, achieves the weighted F1-score of 67.02 (Table I), representing 14.9% increase of the classification accuracy. This comparison highlights the effectiveness of the second LoRA module in improving multi-class classification performance.

#### 2) Visualization of Text Embeddings

We visualize text embeddings to illustrate the motivation for introducing a second LoRA module during fine-tuning. Inspired by the text embedding approach used in BERT [13], which leverages model-generated embeddings for downstream classification tasks, we intend to utilize representations produced by LLM to enhance their performance on multi-class classification problems. As shown in Figure 8, the hidden states from the Qwen1.5-0.5B-Chat model are visualized using the t-SNE dimensionality reduction algorithm on the DR dataset. The visualization indicates that Qwen1.5-0.5B-Chat develops a clear boundary between positive and negative samples, even without pretraining on mental health data. These findings suggest that Qwen1.5-0.5B-Chat has strong inherent representation capabilities. Therefore, fine-tuning a classification head via a second LoRA module on top of these embeddings is a feasible and effective strategy for improving the performance of lightweight models on complex multi-class classification tasks.

**Fig. 8:**
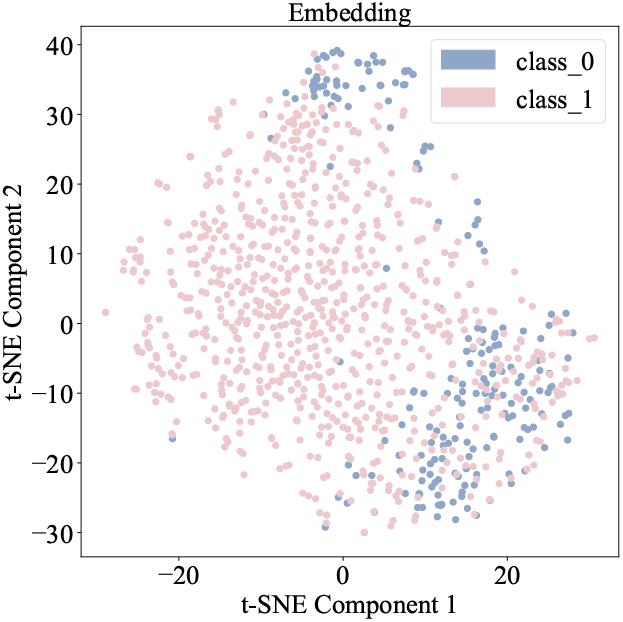
Text Embedding Generated by Qwen1.5-0.5B-Chat.

In the Supplementary Material (Appendix II), we comprehensively compare the text embeddings generated by Qwen1.5-0.5B-Chat, BERT, and the pretrained Mental-BERT. The results show that the performance of Qwen1.5-0.5B-Chat is comparable to that of the pretrained Mental-BERT. In contrast, the standard BERT model fails to establish clear class boundaries, with data points from different classes scattered randomly in the feature space.

### G. Model Efficiency Analysis

This section explores the deployment aspects of the MentalQLM model. First, we compare MentalQLM and MentaLLaMA-7B-Chat in terms of parameter size, GPU memory usage, and inference speed. Next, we examine the impact of LLM quantization methods on classification and reasoning accuracy, providing foundational insights for practical implementation.

#### 1) Computational Efficiency Analysis

All experiments are conducted on a consumer-grade graphics card (RTX 4090). The results in Table IV highlight the significant advantages of MentalQLM over MentaLLaMA-7B-Chat in terms of computational efficiency. We calculate and present the inference time for the test dataset which contains 6,057 samples. Notably, the memory consumption of MentalQLM further underscores its suitability for low-resource environments. It requires only 1.69 GB of GPU memory, compared to 21.27 GB for MentaLLaMA-7B-Chat, making it feasible to be deployed on standard consumer hardware without the need for specialized GPUs.

**TABLE IV:** Computational efficiency between MentalQLM and MentaLLaMA-7B-Chat on the whole test dataset.

| Model | Parameters | GPU Memory | Inference time | vLLM Acceleration | SGLang Acceleration |
| --- | --- | --- | --- | --- | --- |
| MentalQLM | 0.46B | 1.69 GB | 8min | 2min 11s | 5min 24s |
| MentaLLaMA-7B-Chat | 6.74B | 21.27 GB | 137h | 18min 31s | 17min 14s |

In terms of baseline inference speed, the model size of MentaLLaMA-7B-Chat nearly reaches the memory limit of the RTX 4090, which has 24 GB of GPU memory. This limitation prevents the use of large batch sizes and results in an impractical inference time of 137 hours. In contrast, the baseline inference time for MentalQLM is only 8 minutes, representing a substantial improvement in latency. With only 0.46 billion parameters, which is approximately 15 times fewer than MentaLLaMA-7B-Chat’s 6.74 billion, MentalQLM demonstrates exceptional inference speed and resource efficiency. This clear disparity in computational demands highlights the model’s practicality for real-world deployment. The lightweight architecture of MentalQLM enables a single forward-pass inference in just 20 milliseconds, which is a critical metric for latency-sensitive applications such as real-time mental health support.

To further optimize model deployment, we employ the LLM acceleration frameworks vLLM [46] and SGLang [47], which significantly enhance inference speed by optimizing attention mechanisms and enabling high-throughput batched processing. When accelerated using vLLM and SGLang on the multi-threaded setting, MentalQLM reduces inference time to 2 mins 11s and 5 mins 24s, respectively. In comparison, the accelerated inference times for MentaLLaMA-7B-Chat remain considerably slower at 18 mins 31s and 17 mins 14s, despite applying the same optimization techniques.

These metrics collectively validate the deployment feasibility of MentalQLM in resource-constrained scenarios. Its minimal hardware requirements, combined with framework-accelerated performance, effectively address key challenges associated with practical applications. The results emphasize that MentalQLM achieves a strong balance between model capacity and computational efficiency, which is essential for real-world adoption in healthcare settings where accessibility and cost-effectiveness are critical.

#### 2) Quantization Analysis

It is well understood that training LLM in lower precision can optimize computational efficiency, though it may potentially impact model performance. In this section, we evaluate the quantization of Qwen1.5-0.5B-Chat model across different parameter scales. As shown in Table V, we assess the performance of four quantized models under two precision levels: int8 and int4, as well as using the QLoRA-int8 and QLoRA-int4 methods [48].

**TABLE V:**
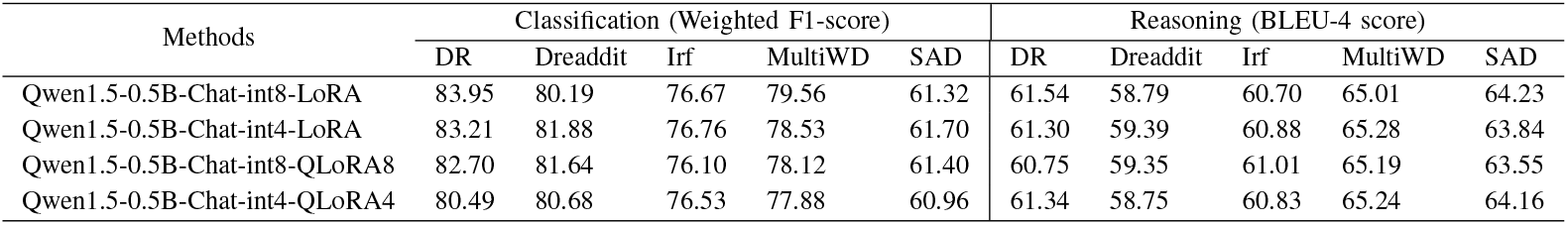
Comparison of LLM model performance with different quantization methods.

The results in Table V demonstrate that even the lightest model, quantized with QLoRA-int4, maintains high classification and reasoning performance. This indicates that quantization does not adversely affect the model ability, and it still performs well on downstream tasks in classification and reasoning. These findings highlight the potential of quantization for developing lightweight and deployable LLM for mental health applications. Such models could be effectively deployed on resource-constrained edge devices without sacrificing performance, paving the way for more accessible mental health support technologies.

## IV. Discussions

In this paper, we propose MentalQLM, a novel lightweight LLM designed for mental disorder classification and diagnostic reasoning. By leveraging a LoRA-enabled parameter-efficient tuning strategy and targeted data selection, knowledge can be effectively transferred to the edge LLM with low computational overhead. We observe that the LLM can generate meaningful embeddings without requiring extensive pretraining on downstream tasks. Building on these embedding, a second LoRA module and a dense classification layer are fine-tuned using multi-class mental health datasets. Experimental results show that our developed MentalQLM achieves strong classification performance and exhibits reasoning capabilities comparable to those of larger-scale models. Nevertheless, we highlight that large LLM still demonstrate superior ability in generating more coherent and contextually rich text.

To this end, we advocate a hybrid approach of developing LLM for enhancing mental healthcare. Lightweight LLM deployed on edge devices (e.g., mobile phones) can be utilized for timely, preliminary diagnostic results and conversational dialogues. While large LLM deployed on cloud platforms can process these conversation records to provide more accurate and detailed suggestions. The predictions and reasoning from large LLM can also benefit fine-tuning the lightweight edge LLM. This dual-model framework enables comprehensive development of mental healthcare systems that delivers both timely and accurate diagnoses, offering patients better insights and treatment recommendations.

The development of lightweight models for mental health-care applications has attracted significant attention in recent years, with notable examples including mhGPT [25] and MentalRoBERTa [49]. Our proposed MentalQLM has clear distinctions from these models. The mhGPT model contains 1.98 billion parameters, approximately four times larger than our MentalQLM. Moreover, our proposed MentalQLM introduces a new dual LoRA strategy and the classification head, and it achieves 14.9% increase in classification accuracy on the SAD dataset (Table I) compared to the single-module approach (Table III) as presented in the mhGPT model. Additionally, MentalRoBERTa [49] is fine-tuned from BERT, which is based on a Transformer encoder architecture and cannot generate text. In contrast, our proposed MentalQLM model enables to produce both classification outputs and textual reasoning, providing interpretable diagnostic explanations.

We acknowledge that the LoRA approach is an established technique for parameter-efficient tuning. Our work introduces a new dual LoRA method and a data pruning strategy, an area that remains largely underexplored. Through comprehensive evaluation across diverse datasets, MentalQLM demonstrates substantial gains in computational efficiency while preserving strong performance in mental disorder classification and reasoning tasks, offering a promising advancement toward bridging the gap between research and practical mental health-care applications. In addition, our ablation analysis of dataset pruning indicated the crucial role of data quality in model fine-tuning. These results indicate that even with limited data, high-quality, well-curated samples are sufficient to maintain strong model performance, whereas noisy or high-perplexity data can negatively impact learning. Furthermore, the model has improved performance on complex multi-class classification tasks by incorporating a second LoRA module along with a classification head. This suggests that the additional capacity and representational flexibility introduced by the second LoRA layer allow the model to better capture nuanced class distinctions, enhancing its overall classification accuracy.

Our future research will focus on enhancing the multi-class classification capabilities of MentalQLM. Although LLMs such as GPT-4 have shown impressive performance on translation tasks, we acknowledge that human evaluation of translation is significant to ensure reliability of the translated text and thus comprehensiveness of the reasoning outputs generated by LLMs. Firstly, for multi-class categorization, ensemble learning can be employed to improve model performance on highly imbalanced datasets [40]. Specifically, distinct classification heads can be trained for head classes and tail classes in the imbalanced dataset respectively, and the results from these classifiers can be aggregated to produce the final output. Secondly, the proposed data pruning method based on perplexity may eliminate rare but clinically relevant examples. Consequently, more advanced data pruning methods can be developed, including linguistic indicators [50] and preference scoring through the reward model based on LLMs [51]. Additionally, to enhance the coherence of reasoning, preference dataset for reasoning can be constructed based on the suggestions of mental health experts, then reinforcement learning algorithms such as Proximal Policy Optimization (PPO) [52] or Direct Policy Optimization (DPO) [53] can be employed to further fine-tune the model’s outputs. These advancements will improve the applicability and effectiveness of lightweight LLM in real-world mental healthcare scenarios. Furthermore, we will validate the effectiveness of our developed MentalQLM in real-world settings, and explore the integration with existing mental healthcare frameworks, including clinical decision support systems, telehealth platforms, and community-based mental health services. Given its compact model size, fast inference speed, and acceleration frameworks, MentalQLM could be deployed on edge devices such as smartphones and wearables. This enables continuous, low-latency monitoring and support without reliance on cloud-based infrastructure. Such capabilities unlock a range of practical applications, particularly in emergency scenarios, such as real-time triage during mental health crises, or just-in-time interventions for individuals experiencing acute anxiety or panic attacks. MentalQLM could also be applied in high stress situations, such as emergency rooms or crisis hotlines by providing risk assessments and therapeutic prompts during initial patient encounters. Through these applications, integrating MentalQLM into mental healthcare workflows holds significant promise for enhancing accessibility, responsiveness, and early detection capabilities in modern mental health systems.

## V. Conclusion

In this study, we proposed MentalQLM, a novel lightweight LLM designed for mental health classification and reasoning, built on the Qwen1.5-0.5B backbone and enhanced through levering a new dual LoRA fine-tuning strategy. Despite its compact size with only 0.5 billion parameters, MentalQLM achieved a 17.7% improvement in accuracy over larger models such as few-shot tuned GPT-4 model, while demonstrating comparable performance in reasoning. These results highlight the potential of our developed lightweight LLM to deliver accurate, efficient, and accessible mental health support, particularly in computationally constrained settings. Our future work will focus on improving multi-class categorization robustness, enhancing the coherence of reasoning outputs, and exploring integration into real-world clinical workflows to broaden the model’s practical impact in mental healthcare.

## Data Availability

All data produced in the present work are contained in the manuscript

