## Supplementary material for "MentalQLM: A lightweight large language model for mental healthcare based on instruction tuning and dual LoRA modules"

### Appendix I
### Datasets and Experiment Setting

In this section, we introduce the datasets used for both monolingual and multilingual evaluation, covering seven languages. We then describe the implementation details, including the selected parameter-efficient tuning methods and the choice of hyperparameters. The evaluation metrics are also presented, including the BLEU-4 for $n$-gram similarity, BERT-score for semantic similarity, BART-score for text probability, and GPT-score for human-like assessment [1], [2].

#### A. Datasets

We used five monolingual IMHI sub-datasets to validate our developed MentalQLM, and we also evaluated the model on multilingual datasets. The datasets DR, dreaddit, Irf, MultiWD, and DEP-SEVERITY were Reddit-sourced data, while the SAD dataset was collected through user interactions. All datasets are publicly available, and the data privacy and user consent have been properly resolved. These datasets have been fully anonymized to ensure that no personal information can be identifiable, which can be categorized as binary-class classification datasets and multi-class classification datasets.

- Binary-class datasets: Four datasets are utilized for binary classification tasks, DR [3], Dreaddit [4], Irf [5], and MultiWD [6]. These datasets focus on determining whether posts show signs of depression, stress, risk factors, or wellness. All datasets are imbalanced, except for Dreaddit, which has a balanced distribution.
- Multi-class dataset: The SAD dataset [7] is used to support multi-classification. This dataset was designed to identify different causes of stress in posts, with categories including school, financial problems, family issues, social relationships, work, health issues, emotional turmoil, everyday decision-making, and other stress causes. The dataset is highly imbalanced, with the last three classes having significantly fewer data points.
- Multilingual dataset: For multilingual evaluation, there are limited open-sourced multilingual datasets that are suitable for mental health study. Instead, as indicated in previous studies [8], [9], they translated English dataset into non-English datasets for multilingual test. Accordingly, we created multilingual datasets for mental health tests following their recommended pipeline. Specifically, we utilized LLM (GPT-4o mini) approach translating the DEP-SEVERITY dataset into six different languages (Turkish, French, Portuguese, German, Greek, and Finnish). DEP-SEVERITY is an imbalanced multi-class classification dataset designed to assess users' depression levels, categorized as minimum, mild, moderate, or severe. The dataset comprises a total of 3,553 samples and the mean length of the tokenized input sentence is 189.

Details of the six datasets for monolingual and multilingual evaluation are presented in Table I.

#### B. Implementation Details

The Qwen1.5-0.5B-Chat is chosen as the backbone LLM model, which is trained on a single 4090 GPU with the LLaMA-factory framework [11], [12]. There are different parameter-efficient fine-tuning (PEFT) methods in literature, such as adapters, prefix tuning, prompt tuning, and LoRA [13], [14]. These studies comprehensively evaluated and compared LoRA approach with these PEFT methods. They explored the theoretical foundations of LoRA, and their results showed that the LoRA method significantly reduces the number of trainable parameters while maintaining high performance, representing a significant step forward in making LLM fine-tuning more accessible, scalable, and cost-effective. Therefore, this paper used the LoRA approach for model fine-tuning.

We developed a dual-LoRA module for the fine-tuning. For hyperparameter selection of the first LoRA module, prior research suggests that a low-rank adaptation matrix is sufficient, as increasing the LoRA rank $r$ does not capture a more meaningful subspace [14]. Moreover, a higher rank $r$ would introduce additional computational costs, conflicting with the lightweight design of MentalQLM. Accordingly, we adopt a smaller LoRA rank of 8, consistent with earlier work on fine-tuning LLMs for mental health conversations [15]. The LoRA scaling coefficient $\alpha$, which defines the influence of matrix $\mathbf{B}$ on matrix $\mathbf{A}$, is set to $\alpha = 2r$ to ensure gradient stability and approximate full parameter fine-tuning performance [16]. Based on the sentence lengths in the instruction-tuning dataset, we set the maximum input and output lengths to 384 under

TABLE I: Summary of datasets for mental health instruction tuning.

| Datasets | Task | Data Size (Train / Val / Test) | Class Proportion | Instruction Length | Output Length |
|---|---|---|---|---|---|
| DR [3] | Binary depression classification | 1,003 / 430 / 405 | 79.9% / 20.1% | 251 | 120 |
| Dreaddit [4] | Binary stress classification | 2,837 / 300 / 414 | 52.7% / 47.3% | 116 | 95 |
| Irf [5] | Binary risk factors classification | 3,943 / 985 / 2,113 | 63.6% / 36.4% | 164 | 135 |
| MultiWD [6] | Binary wellness classification | 15,744 / 1,500 / 2,441 | 36.8% / 63.2% | 167 | 79 |
| SAD [7] | Multi-class stress cause classification | 5,547 / 616 / 684 | 24.7% / 18.6% / 12.3%<br>10.3% / 9.6% / 8.6%<br>6.4% / 6.3% / 3.1% | 31 | 78 |
| DEP-SEVERITY [10] | Multi-class depression assessment | 2,842 / 355 / 356 | 72.8% / 8.2%<br>11.1% / 7.9% | 189 | / |

a bf-16 precision setting. For hyperparameter selection of the second LoRA module, since this training stage is task-specific and involves fine-tuning based on a single task, which is simpler compared to language regression modeling in the first stage, and according to previous research [14], the LoRA rank $r$ is set to 4 to reduce the parameter count. Table II lists details of selected hyperparameters for the two LoRA modules.

For the training process, we adopt the default configurations recommended by the LLaMA-Factory training framework [12], which could make it easier for researchers to reproduce our work. In the instruction tuning stage, all the models are trained for 15 epochs. The checkpoint of the last step is chosen to output results on the test set. The model is trained with the AdamW optimizer [17], and we set the maximal learning rate of 5e-5 with the cosine decay learning rate regulator. In the fine-tuning stage, the model is trained for 30 epochs, and the model achieving the highest classification accuracy on the validation set is saved for evaluation.

TABLE II: Network configurations for MentalQLM.

| Parameters | Settings |
|---|---|
| **Stage #1 Instruction Tuning** | |
| LoRA I rank ($r$) | 8 |
| LoRA I scale factor ($\alpha$) | 16 |
| Max input length | 512 |
| Max output length | 512 |
| Tunable parameters | 3,784,704 |
| **Stage #2 Multi-class Fine-tuning** | |
| Max input length (SAD) | 32 |
| Max input length (DEP-SEVERITY) | 256 |
| LoRA II rank ($r$) | 4 |
| LoRA II scale factor ($\alpha$) | 8 |
| Tunable parameters | 393,216 |

#### C. Evaluation Metrics

Four metrics are used to evaluate the classification and reasoning performance of our proposed MentalQLM. For a fair comparison with MentaLLaMA [18], we used the recommended weighted F1 score to evaluate all classifiers. The weighted F1-score is a metric that combines precision ($P$) and recall ($R$) into a single balanced measure, calculated as their harmonic mean. For multi-class classification tasks, we compute the F1-score for each class and average them, weighted by the number of instances in each class, which can be formalized as

$$\text{F1\_weight} = \sum_{i=1}^{n} \left( \frac{2 \cdot P_i \cdot R_i}{P_i + R_i} \right) \cdot \frac{S_i}{\sum_{j=1}^{n} S_j} \cdot 100 \tag{1}$$

where $n$ is the number of classes, $P_i$ and $R_i$ represent the precision and recall of class $i$ respectively, and $S_i$ denotes number of instances of class $i$.

The BLEU (Bilingual Evaluation Understudy) score is a metric used to evaluate the quality of machine-generated text. Specifically, BLEU-4 score focuses on comparing $n$-grams up to 4-grams between the candidate translation and the reference, which can be defined as

$$\text{BLEU-4} = BP \cdot \exp \left( \sum_{n=1}^{4} \frac{w_n \cdot clip_n}{total_n} \right) \cdot 100 \tag{2}$$

where $BP$ is the brevity penalty that penalizes candidates shorter than the reference, $w_n$ are weights with $\sum_{n=1}^{4} w_n = 1$, typically $w_n = \frac{1}{4}$ for BLEU-4, $clip_n$ is the clipped count of $n$-grams matching the references, and $total_n$ is the total count of n-grams in the candidate translation. The BLEU-4 score effectively measures the similarity between machine-generated text and reference answers.

In addition to BLEU-4 score, the BERT-score [19] and BART-score [20] are introduced to evaluate the reasoning ability of different LLMs. The BERT-score evaluates text generation quality by measuring semantic similarity between candidate and reference texts using contextualized embeddings from the BERT model [19]. It computes token-wise cosine similarities between the embedding, ensuring alignment based on meaning rather than exact word matching. BERT-score can be calculated as the harmonic mean of precision and recall:

$$P = \frac{1}{|\mathbf{c}|} \sum_{i} \max_{j} \text{cossim}(\mathbf{c}_i, \mathbf{r}_j) \tag{3}$$

$$R = \frac{1}{|\mathbf{r}|} \sum_{j} \max_{i} \text{cossim}(\mathbf{r}_j, \mathbf{c}_i) \tag{4}$$

$$\text{BERT-score} = \frac{2PR}{P + R} \tag{5}$$

where $\mathbf{c}_i$ denotes contextual embedding vector for the $i$-th token in the candidate text, $\mathbf{r}_j$ denotes contextual embedding vector for the $j$-th token in the reference text.

In contrast, the BART-score leverages the autoregressive nature of the BART model to assess generation quality through sequence-level probabilities, reflecting how likely the generated text aligns with the reference [20], which can be formalized as:

$$\text{BART-score} = -\frac{1}{|c|} \sum_{t=1}^{|c|} \log P(c_t | c_{<t}, r) \tag{6}$$

where $c$ denotes the candidate text, $r$ denotes the reference text, $P(c_t|c_{<t}, \mathbf{r})$ denotes given previous text $c_{<t}$ and reference text $r$, the probability of generating the next token $c_t$.

By using these four metrics, we aim to comprehensively evaluate the generated text, capturing both semantic similarity and probabilistic coherence, therefore offering a robust assessment of reasoning quality.

### APPENDIX II
### VISUALIZATIONS OF TEXT EMBEDDINGS

Inspired by the text-embedding model BERT [21], which utilizes embeddings generated by the model for downstream classification tasks, we aim to leverage representations produced by LLM to enhance their performance on multi-class classification tasks. As shown in Figure 1, the hidden states from Qwen1.5-0.5B-Chat, BERT, and the pre-trained Mental-BERT are visualized using the t-SNE dimensionality reduction algorithm. This visualization is performed on two binary classification datasets, DR and Irf. In bidirectional text embedding models such as BERT and Mental-BERT, the embedding of the [*CLS*] token is visualized, because it is designed to capture aggregated semantic information from the entire input sentence. This token serves as a standard representation for sentence-level embeddings in such architectures.

The results demonstrate that the Qwen1.5-0.5B-Chat develops a distinct boundary between positive and negative samples, even without fine-tuning on mental health datasets. Its performance is comparable to that of the pre-trained Mental-BERT model. In contrast, the standard BERT model fails to establish a clear boundary between classes, with points of different classes scattered randomly in the feature space. These findings suggest that the Qwen1.5-0.5B-Chat has robust inherent representation capabilities comparable to these domain-specific pre-trained models, without requiring specialized pre-training. Consequently, it is feasible to finetune a classification head with a second LoRA module based on these embeddings.

### APPENDIX III
### IMBALANCE DATA PROCESSING

The SAD and DEP-SEVERITY datasets are highly imbalanced. For example, the least represented classes in these datasets account for only 3.1% and 7.9% of the total samples, respectively. To mitigate this issue, we employed the up-sampling technique to increase the number of samples in the underrepresented classes, matching them to the size of the majority class. Specifically, following the approach in previous study [22], we used the random over-sampling (ROS) approach, which randomly duplicates samples from the minority classes to re-balance the dataset before training.

As shown in Table III, this up-sampling strategy effectively mitigated the data imbalance issue on both the SAD and DEP-SEVERITY datasets. In particular, the model achieved higher F1-scores on seven out of eight datasets after applying the up-sampling method, with the largest improvement as 29.6% increase on the DEP-SEVERITY-PT dataset.

### APPENDIX IV
### PROMPT FOR EVALUATION

The prompt used to rate the generated sentences is listed in Table IV. The sentences are evaluated based on helpfulness, relevance, accuracy, and detailedness [2]. Each of these aspects is rated on a scale from 1 to 10, ensuring comprehensive feedback with reduced bias.

TABLE III: Performance of the imbalance training techniques.

| Dataset | Precision | Recall | F1 |
|---|---|---|---|
| | *Before up-sampling* | | |
| SAD | 57.95 | 56.02 | 55.67 |
| SEP-SEVERITY-EN | 55.28 | 56.48 | 55.46 |
| DEP-SEVERITY-TR | 48.76 | 38.87 | 41.55 |
| DEP-SEVERITY-FR | 52.87 | 46.41 | 48.81 |
| DEP-SEVERITY-PT | 34.67 | 33.27 | 33.06 |
| SEP-SEVERITY-DE | 46.11 | 39.13 | 40.52 |
| DEP-SEVERITY-GR | 45.55 | 30.95 | **31.62** |
| DEP-SEVERITY-FI | 32.14 | 31.90 | 31.87 |
| | *After up-sampling* | | |
| SAD | 57.25 | 61.94 | **57.28** |
| SEP-SEVERITY-EN | 60.04 | 54.12 | **56.64** |
| DEF-SEVERITY-TR | 45.46 | 41.57 | **43.05** |
| DEF-SEVERITY-FR | 56.30 | 52.62 | **53.48** |
| DEP-SEVERITY-PT | 42.74 | 43.72 | **42.84** |
| DEP-SEVERITY-DE | 42.82 | 44.17 | **42.89** |
| DEP-SEVERITY-GR | 29.67 | 29.78 | 29.42 |
| DEP-SEVERITY-FI | 36.12 | 33.98 | **34.70** |

*EN, TR, FR, PT, DE, GR, FI stand for English, Turkish, French, Portuguese, German, Greek and Finnish, respectively.

TABLE IV: Prompt for reasoning evaluation.

| **Prompt for reasoning ability evaluation.** |
|---|
| **System prompt**<br>You are a mental health expert for checking the quality of the answer.<br><br>**User prompt**<br>[Question]<br>[The Start of Assistant 1's Answer]<br>{Text}<br>[The End of Assistant 1's Answer]<br>[The Start of Assistant 2's Answer]<br>{Text}<br>[The End of Assistant 2's Answer]<br><br>We would like to request your feedback on the performance of two AI assistants in response to the user question displayed above.<br>Please rate the helpfulness, relevance, accuracy, level of details of their responses. Each assistant receives an overall score on a scale of 1 to 10, where a higher score indicates better overall performance.nPlease first output a single line containing only two values indicating the scores for Assistant 1 and 2, respectively.<br>The two scores are separated by a space. In the subsequent line, please provide a comprehensive explanation of your evaluation, avoiding any potential bias and ensuring that the order in which the responses were presented does not affect your judgment. |

### APPENDIX V
### EXAMPLES OF GENERATED NON-ENGLISH CHARACTERS

As shown in Table V, in the exploration of reasoning outputs of MentalQLM, we observed a small portion of non-English characters in the generated text. The reason is that most of Qwen's training corpus is in Chinese, so fine-tuning the LLM

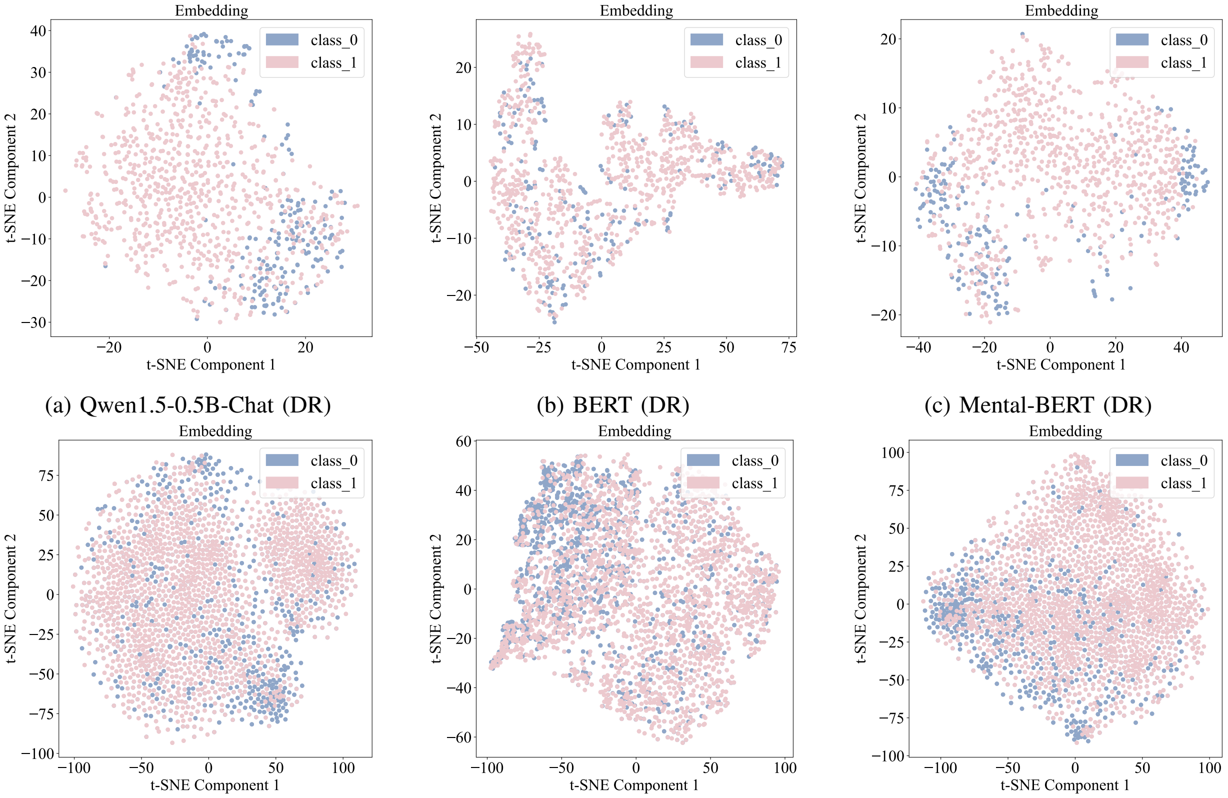


(a) Qwen1.5-0.5B-Chat (DR) (b) BERT (DR) (c) Mental-BERT (DR)

(d) Qwen1.5-0.5B-Chat (Irf) (e) BERT (Irf) (f) Mental-BERT (Irf)

Fig. 1: Visualizations of embeddings for Qwen1.5-0.5B-Chat (representing LLM), BERT (representing traditional embedding models), and Mental-BERT (representing pre-trained embedding models). (a)-(c) depict embeddings on the DR dataset, (d)-(e) demonstrate embeddings on the Irf dataset.

using a dataset that aligns with the distribution of the training corpus will yield more reliable results.

TABLE V: Examples of generated non-English characters.

| **Non-English characters during reasoning.** |
|---|
| **'Output1'**:'No, the poster does not suffer from depression. Reasoning: The post does not exhibit any emotional indicators of depression. It simply mentions having new ven 蚀 ualan friends at a local restaurant. There is no mention of emotional distress, feelings of sadness, hopelessness, or any other symptoms commonly associated with depression.' |
| **'Output2'**: No, the post does not show this risk. Reasoning: In this post, the individual is discussing a topic that is not explicitly mentioned but is being used as a metaphor for a common theme of suicide. While the post mentions social awkwardness and the use of clichés, it does not indicate any clear signs of thwarted belongingness or inadequate connectedness. The individual's focus is on the idea of suicide, rather than their own feelings of loneliness or lack of social support. |
| **'Output3'**: No, the stress in this post does not belong to the specific category of School, Financial problems, Family issues, 和社会 relationships. The post does not provide any specific information about the cause of stress. It is possible that the stress is related to work or career issues, but without further context, it is not possible to determine the exact cause. |

### Appendix VI
### Examples of high perplexity pairs

As shown in Table VI, it is noted that some inputs are fairly short, such as “friend OK No” and “Please help my friend”. While other posts are ambiguous for mental disorder, making it challenging to determine whether they indicate symptoms of disorders.

TABLE VI: Examples of high perplexity pairs.

| **Instruction-output pairs with high perplexity.** |
|---|
| **'instruct'**: 'Consider this post: ”Why did I open my mouth I should've just said “I'm fine” Ĩ don't need help Or maybe I do” Question: Does the poster suffer from stress?' **'output'**: 'Yes, the poster suffers from stress. Reasoning: The poster expresses regret and indecision about opening up and asking for help. This suggests internal conflict and possible stress. The usage of phrases like ”should've just said 'I'm fine'” and ”Or maybe I do” indicates a struggle with vulnerability and a need for support. Overall, the language used implies that the poster may be experiencing stress.' |
| **'instruct'**: 'Consider this post: ”friendship ok No” Question: Does the poster suffer from depression?' **'output'**: 'No, the poster does not suffer from depression. Reasoning: The post lacks sufficient information to determine if the poster suffers from depression. It only consists of two words and does not provide any context or emotional indicators.' |
| **'instruct'**:'Consider this post: ”I feel like no matter how hard I try when I make one step forward I,am always two steps back. it,is so defeating and embarrassing Question: Does the post show risk of perceived burdensomeness?” **'output'**:'No, the post does not show this risk. Reasoning: 1. The post mentions feelings of defeat and embarrassment, but does not directly indicate thoughts of burden or being better off gone. These feelings suggest a sense of frustration and disappointment, but not necessarily perceived burdensomeness.The post does not mention any social or interpersonal difficulties that could contribute to feelings of perceived burdensomeness. It focuses more on personal setbacks and struggles. The response also does not indicate any perceived burdensomeness.' |

### APPENDIX VII
### EXAMPLE OF GENERATED REASONING TEXT

Inspired by previous studies [2], [23]–[25], GPT-4o is utilized to evaluate the quality of generated sentences by MentalQLM and MentaLLaMA-7B-Chat. Table VII presents the sentences generated by these models.

TABLE VII: Example of generated reasoning sentences by different LLMs.

| Instruction | Model | Score | Output |
|---|---|---|---|
| Consider this post: ”The one and only thing I want to and have a will to do is to listen to the music all day and space out in it. Everything else is boring, draining and dull. And I receive no mental pleasure from doing anything else.” Question: Does the poster suffer from depression? | MentalQLM-0.5B | 7 | Yes, the poster suffers from depression. Reasoning: The post expresses a desire to isolate oneself and avoid social interaction. The poster describes music as a source of pleasure and relaxation, indicating a potential desire for escape from negative emotions and a lack of enjoyment in other activities. The mention of receiving no mental pleasure from doing anything else suggests a possible disconnection from their own emotions and a lack of fulfillment in their life. These feelings align with symptoms commonly associated with depression, particularly the desire to withdraw from social interactions and a lack of enjoyment in activities. |
| | MentaLLaMA-7B-Chat | 8 | Yes, the poster suffers from depression. Reasoning: The post conveys a strong sense of emotional numbness and a lack of interest or enjoyment in activities other than listening to music. The poster describes everything else as boring, draining, and dull, indicating a lack of fulfillment or satisfaction in their daily life. This suggests a possible symptom of depression, as individuals with depression often experience a loss of interest or pleasure in activities they once enjoyed. The mention of receiving no mental pleasure from doing anything else further emphasizes the emotional emptiness and dissatisfaction commonly associated with depression. |
| | GPT-4o (Judge) | / | Both AI assistants provided helpful, relevant, and accurate responses with a decent level of detail. They were able to identify the potential symptoms of depression in the post and provide reasoning for their conclusions. Assistant 2’s response was slightly more detailed and emphasized the emotional emptiness and dissatisfaction commonly associated with depression, which was a valuable addition to their analysis. However, both assistants could have provided more specific examples to support their reasoning. Overall, I found their responses to be informative and insightful. |
| Consider this post: ”The chapter we shared together may have ended but the story itself will never change. So here’s to you, Babu. Thank you for teaching me so much, for the irreplaceable memories, and for being such an important part of my life. — **tl;dr**: I love my current partner, but I think I’ll love my first love forever.” Question: Does the poster suffer from stress? | MentalQLM-0.5B | 8 | No, the poster does not suffer from stress. Reasoning: The poster expresses gratitude and love for their current partner and their past relationship. While the post reflects a sense of nostalgia and longing, there is no indication of emotional distress or stress. The language used is positive and focused on the positive aspects of their relationship, rather than expressing negative emotions or struggles. Therefore, the poster does not appear to be suffering from stress based on this post. |
| | MentaLLaMA-7B-Chat | 7 | No, the poster does not suffer from stress. Reasoning: The poster expresses love and gratitude towards their current partner, but also mentions their first love. While they may have mixed feelings about their past relationship, there is no indication of emotional distress or anxiety in the post. The language used suggests a reflection on past experiences rather than current stress. |
| | GPT-4o (Judge) | / | Both Assistant 1 and Assistant 2 provided relevant and accurate responses to the user’s question. They correctly identified that the poster in the given scenario does not appear to be suffering from stress based on the content of the post. Assistant 1’s response scored slightly higher due to its slightly more detailed analysis of the language. |
| Consider this post: ”my grades are so bad. i am the only one who failed out of the grade in my class and now im all alone in a new class full of failures like me and i really dont know what to do. it feels like all my friends from my previous class are just subtly looking down on me, and i hate it. Fuck the education system and fuck myself for not being able to pass.” Question: Does the spiritual wellness dimension exist in the post? | MentalQLM-0.5B | 8 | No, this wellness dimension does not exist in the post. Reasoning: In the post, the individual expresses feelings of frustration and self-criticism, but there is no mention of a search for meaning or purpose in human existence. The focus is on their academic struggles and feelings of isolation, rather than a broader existential search. |
| | MentaLLaMA-7B-Chat | 9 | No, this wellness dimension does not exist in the post. Reasoning: In the post, the individual expresses feelings of frustration and self-criticism regarding their grades and academic performance. While they may be experiencing emotional distress, there is no mention of a search for meaning or purpose in human existence. The focus is primarily on their academic struggles and negative self-perception. Therefore, the post does not reflect the wellness dimension of the spiritual aspect according to Dunn’s model. |
| | GPT-4o (Judge) | / | Both Assistant 1 and Assistant 2 provided clear and accurate responses to the question about the existence of the spiritual wellness dimension in the post. They both correctly identified that this dimension was not present based on the individual’s focus on academic struggles and self-criticism rather than a search for meaning or purpose in human existence. Both responses were relevant to the user’s question and provided a good level of detail in explaining their reasoning. Assistant 2 received a slightly higher score of 9 due to the additional mention of Dunn’s model, which added depth to the analysis. Overall, both assistants performed well in addressing the user query effectively. |
